# Real-life observation of wildfire-smoke impaired COVID-19 vaccine immunity

**DOI:** 10.1101/2024.10.25.24316123

**Authors:** Gursharan Kaur Sanghar, Melissa Teuber, Resmi Ravindran, Emma Jean Keller, Sean Raffuse, Pedro Hernandez, Angela Linderholm, Gabrielle Echt, Lisa Franzi, Kaelyn Tuermer-Lee, Maya Juarez, Timothy Albertson, Imran Khan, Angela Haczku

## Abstract

**Background:** Wildfires are increasingly common with wildfire smoke affecting millions globally, yet its impact on immune responses is poorly understood. Natural Killer (NK) cells play a role in mediating air pollutant effects and regulating vaccine immunity.

**Objective:** This real-world study, conducted on participants in the Pfizer BNT162b2 COVID-19 vaccine trial, studied the effects of wildfire smoke exposure on long-term vaccine effects.

**Methods:** We collected blood samples from 52 healthy, non-smoking participants (ages 26–83) before and 1 month after placebo or vaccine injections during heavy wildfire smoke events in Sacramento. The study included 28 vaccinated (Group 1) and 24 placebo-injected (Group 2) individuals, the latter vaccinated several months later, outside wildfire season. Blood samples from both Group 1 and 2 were also investigated 6 months after the second dose of vaccine. We analyzed intracellular cytokines, B and NK cell markers by flow cytometry, and serum immunoglobulin levels against common coronaviruses using multiplex assays.

**Results:** A robust S-RBD-specific IgG response observed 1 month post booster, declined variably 6 months later. Wildfire smoke acutely increased IL-13 expression by CD56^bright^ NK cells. IL-13^+^CD56^bright^ NK cells at the time of vaccination negatively correlated with anti-S-RBD IgG (r=-0.41, p<0.05) one month later. Total IgG levels on the other hand, positively correlated with the air quality index (AQI) measured during vaccination (r=0.96, p<0.01). Similarly to age (but not sex, BMI or race/ethnicity), the two-week AQI averages during vaccination showed a significant negative correlation with anti-S-RBD IgG levels 6 months later (r=-0.41, p<0.05).

**Conclusion:** Wildfire smoke may lead to inappropriate immunoglobulin production and diminished vaccine immunity. Our novel findings highlight a previously unrecognized pathway involving NK-cell derived IL-13 and non-specific B-cell activation and underscore the significance of environmental exposures in shaping immunity.

**Clinical Trial registration:** Not applicable

**Key Messages:**

- Wildfire smoke exposure acutely increased IL-13 expression in NK cells, which in turn negatively correlated with COVID-19 vaccine immunity.
- Higher AQI and age during vaccination was negatively associated with anti-SARS-CoV-2 IgG levels six months later.
- Environmental factors such as air pollution caused by wildfire smoke can significantly impact vaccine efficacy through immune modulation.

**Capsule Summary:** Wildfire smoke exposure impairs long-term COVID-19 vaccine immunity potentially through increased IL-13 expression in NK cells.

**Graphical abstract.**
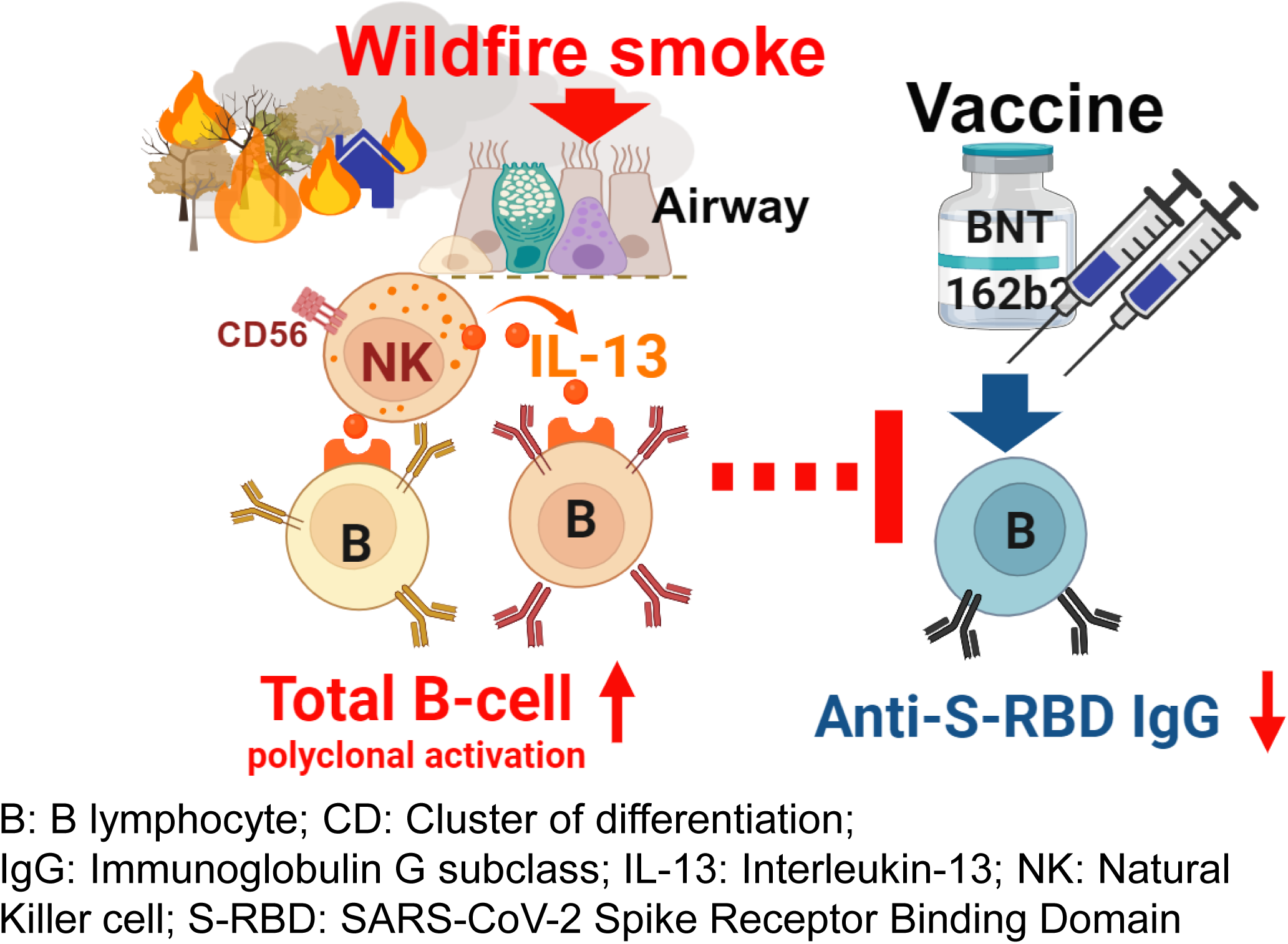
Wildfire smoke during BNT162b2 COVID-19 vaccination induced IL-13^+^CD56^bright^NK cells, increased B cell count and total IgG and decreased antigen-specific IgG

## Introduction

Wildfires are becoming larger, longer lasting, and more intense, with smoke affecting millions of people worldwide^1^. While the acute impacts of wildfire smoke on the respiratory and cardiovascular systems are well documented^2^, the long-term effects are less studied and require further investigation to inform effective mitigation and prevention strategies^3^. Notably, the influence of wildfire smoke on immune responses remains particularly poorly understood.

Exposure to air pollutants has been shown to adversely affect immune cell function, potentially impacting the efficacy of immunizations. Recent studies reported that individuals exposed to higher levels of air pollution exhibited a reduced antibody response to COVID-19 vaccines, suggesting that air pollutants may diminish vaccine-induced immunity^4, 5^. Additionally, long-term exposure to particulate air pollution demonstrated to weaken immune cells in the lungs, potentially undermining the body’s defense mechanisms against respiratory infections and affecting responses to immunizations^6^.

Natural Killer (NK) cells are innate immune cells found throughout the body including in the respiratory tract mucosal tissue, that play an important role in immune responses to viral infections^7–9^. We previously reported in a mouse model that lung NK cells were responsible for promoting dendritic cell lymph node homing, an essential process in developing protective immunity. NK cells also mediated the effects of the toxic air pollutant, ozone (O3)^10^. Other studies suggested that NK cell activation during BNT162b2 vaccination may reduce durability of vaccine-induced antibody responses^11, 12^.

We conducted a real-world study on UC Davis participants of the Pfizer BNT162b2 vaccine trial^13^, that allowed us the unique opportunity to investigate how exposure to wildfire smoke during vaccination may affect NK cell function and long-term vaccine immunity.

## Results and discussion

52 healthy subjects from the Sacramento region (26-83 years old) who were recruited in the original Pfizer BioNTech BNT162b2 vaccine trial^13^ at UC Davis Medical Center in August 2020 consented to enroll in our study. Because of the real-world nature of our study, we were originally blinded to the subjects’ group assignments. The vaccination and blood collection timeline with the daily air quality index (AQI) for fine particulate matter (PM2.5) and O3 are illustrated in **Figure 1A**. Although AQI serves as a proxy for smoke exposure, it offers an accessible and relevant measure for assessing the potential impact of environmental factors on vaccination outcomes. Our study subjects came from the Sacramento County area where AQI monitoring stations in different locations showed similar level of pollution (**Figure 1E**). To be able to correlate exposure levels to immune responses, in this study we integrated the daily AQI data for PM2.5 and O3 levels (**Figure 1B**).

**Figure 1.**
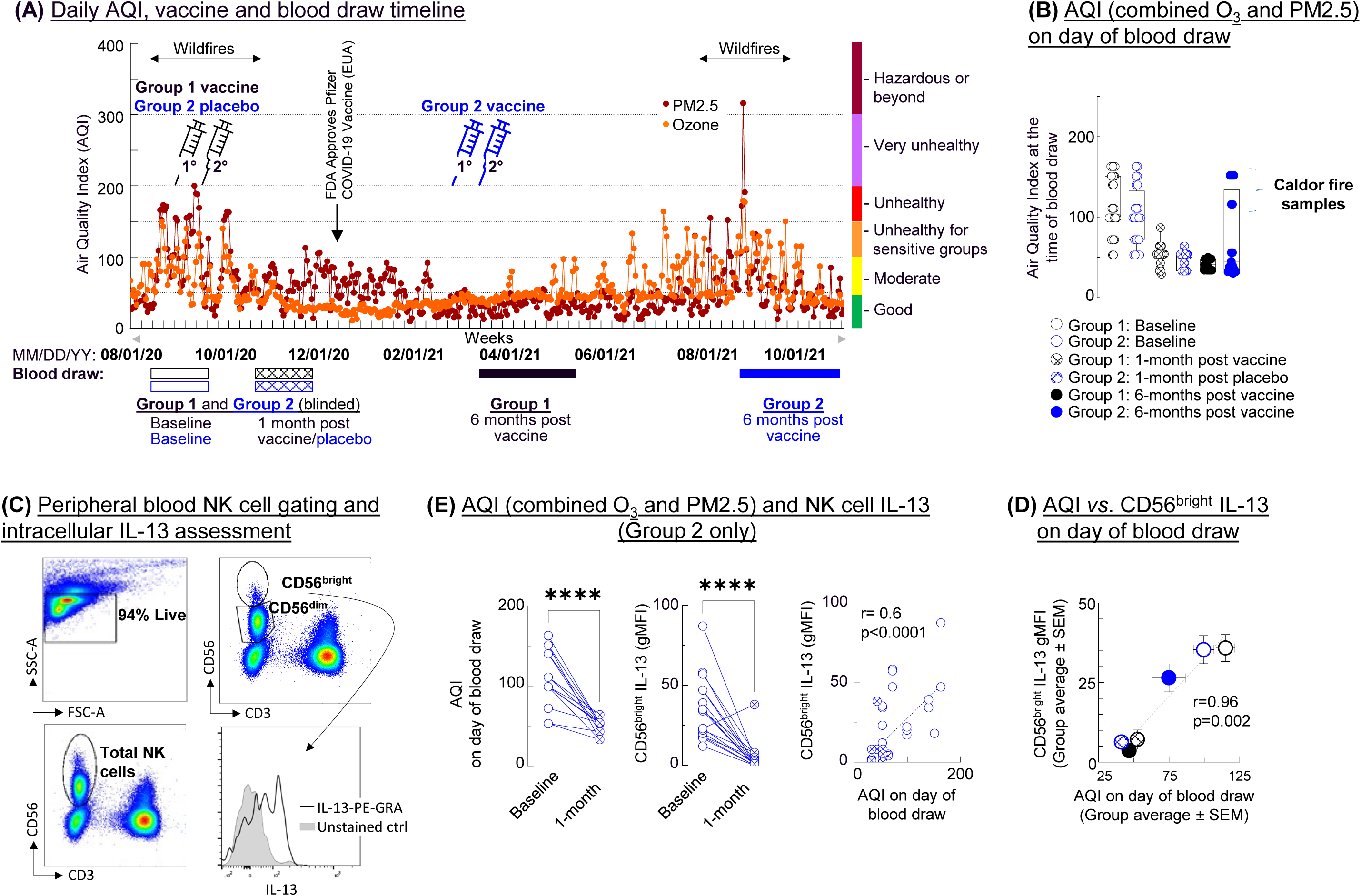
Wildfire smoke exposure increased circulating NK cell expression of IL-13. **(A)**: Vaccination and blood collection timeline with daily air quality index (AQI) for fine particulate matter (PM2.5, dark red) and O_3_ (orange). **(B)** Blood samples were drawn from Group 1 and Group 2 at baseline; 1 month after vaccination (Group 1) or placebo injection (Group 2) and 6 months post vaccination. Bracket indicates the Group 2 (6 months post) samples collected during the Caldor Fire that were removed from immunoglobulin assessment. **(C):** CD56bright and CD56dim NK cells were gated and **(D):** assessed for IL-13 expression. **(E):** Group averages (Mean±SEM) of CD56bright IL-13 *vs.* AQI on day of blood draw (Pearson correlation)

To monitor the dynamics of immune responses and investigate both short-term and long-term vaccine- induced immunity, blood samples were collected at three key time points (baseline, 1 month, and 6 months post-vaccination). Baseline blood samples were collected from all subjects. Group 1 received the initial (1°) and booster (2°) vaccine shots (30µg of the BNT162b2 vaccine) 3 weeks apart. Group 2 received placebo (saline) during the same schedule, all under heavy wildfire smoke conditions. The second blood sample was collected 1 month after the second dose of vaccine (Group 1) or placebo (Group 2) under clean air conditions. Once FDA approved the BNT162b2 vaccine (on December 11, 2020), the group identities of the Pfizer vaccine trial volunteers were revealed. Our study included 28 vaccinated (Group 1, black symbols throughout) and 24 placebo-injected (Group 2, blue symbols) individuals. This latter group then received their vaccine several months later, outside of wildfire season, in March-April 2021 (**Figure 1A-B**). This design allowed us to compare immunoglobulin levels 6 months post vaccination between Group 1 that received the vaccine during wildfires and group 2 that received it under clean air conditions. In 6 subjects in Group 2 however, the 6 months-post vaccine blood draws occurred during the 2021 Caldor wildfires (under extreme AQI conditions indicated by the bracket in **Figure 1B**). To ensure accurate comparisons and minimize confounding effects, these samples were excluded from 6-month immunoglobulin assessments (please, see additional details of Methods in the Online repository).

NK cell activation is involved in the effects of environmental exposures in mice^10, 14^ and in wildfire smoke exposure in a recent study on human subjects^15^. IL-13, a major inflammatory cytokine and activated NK cell product^14^ modulates both innate and adaptive immune responses, and can affect B cell activation and immunoglobulin production. We studied CD56^bright^ and CD56^dim^ NK cell subtypes because of their differential functional capabilities. CD56^bright^ cells (only present in a small proportion [<10%] in peripheral blood, **Figure 1C**), are major producers of pro-inflammatory cytokines^7^ but their role in IL-13 production especially in response to wildfire smoke has not been described before. While we found no changes in IL-13 expression in CD56^dim^ NK cells, CD56^bright^ NK cells showed highly significantly greater IL-13 expression in samples drawn during wildfire smoke events (**Figure 1D**). In fact, the AQI levels on the day of blood draw showed a strong positive correlation in the individual Group 2 samples (**Figure 1D**, right panel) and across all 6 group averages (**Figure 1E**). These results demonstrate that exposure to wildfire smoke is associated with increased IL-13 production from CD56^bright^ NK cells suggesting their potential to impact vaccine-induced immunity.

We observed a robust induction of S-RBD-specific IgG one month after the booster dose in vaccinated individuals (Group 1), while placebo-injected subjects (Group 2) showed no detectable response at the same time point (**Figure 2A**). Although the 1-month S-RBD-specific IgG levels significantly correlated with 6-month levels (**Figure 2B**), the latter showed substantial variability among subjects. Notably, there was significant cross-reactivity between anti-SARS-CoV and anti-SARS-CoV-2 S-RBD IgG (**Figure 2C**), while vaccination did not induce antibodies against MERS-CoV S-RBD, nucleocapsid (N), or total IgG (**Figure E2A**). IgG1 was the predominant isotype among anti-S-RBD antibodies (**Figure 2D**). As expected, age was inversely correlated with SARS-CoV-2 S-RBD IgG levels at both 1- and 6-months post-vaccination (**Figure 2E**). Importantly, there were no statistical differences in age, sex, race/ethnicity, or body mass index between Groups 1 and 2, ensuring that AQI (wildfire smoke exposure) during vaccination remained the main distinguishing factor (**Figure 2F; Figure E3**).

**Figure 2.**
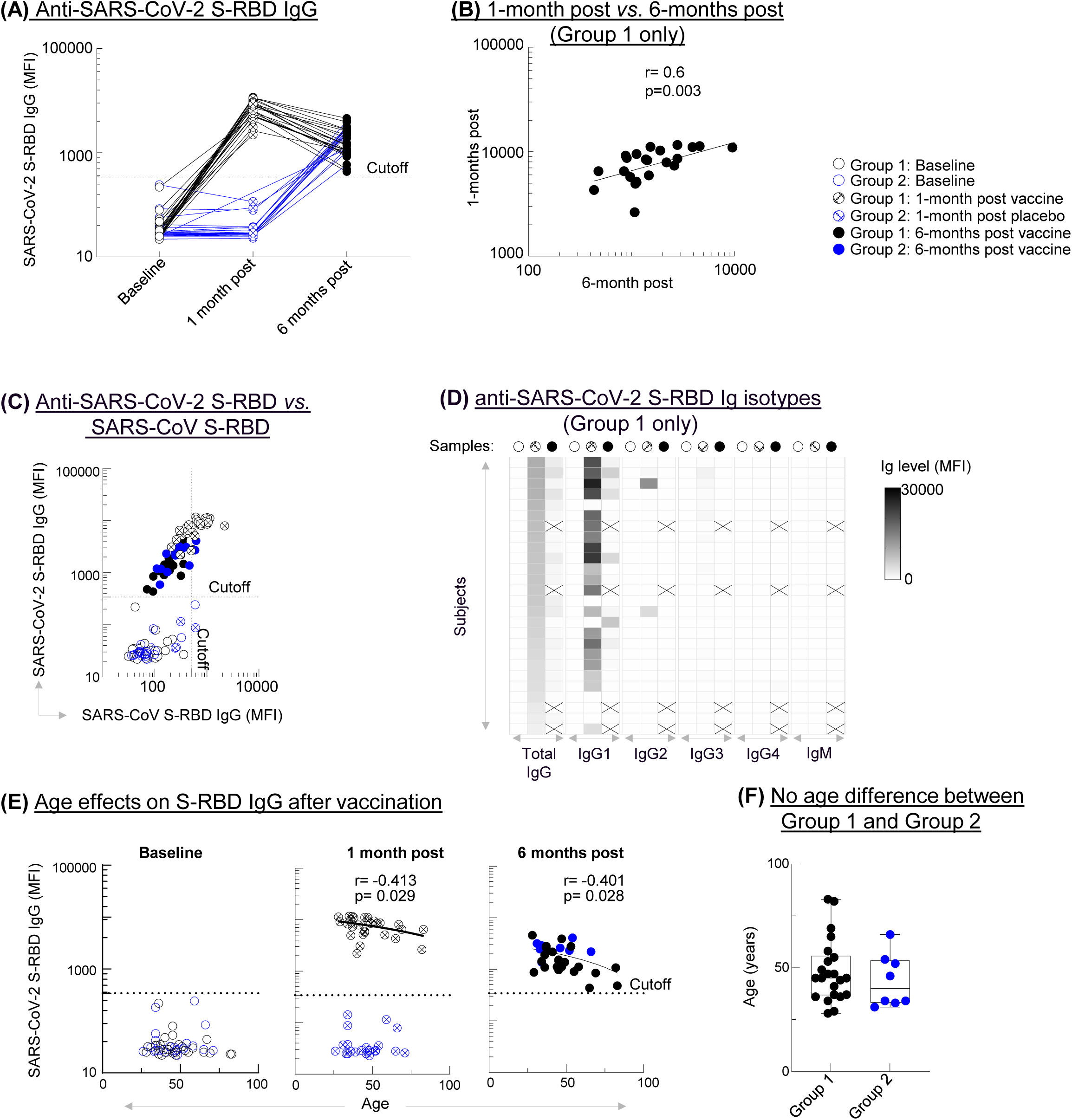
Anti-SARS-CoV-2 S-RBD response to BNT162b2 COVID-19 vaccination was reduced by age. **(A):** Anti-SARS-CoV-2 S-RBD IgG levels (LUMINEX) in Group 1 (black) and Group 2 (blue, originally placebo) samples at baseline [open circles], 1 month-post vaccine (Group 1) or placebo (Group 2), [crossed circles] and 6-month post vaccine [full circles] **(B):** Correlation between 1- and 6-months post vaccine in Group 1 (Pearson correlation). **(C):** Cross reactivity between anti-SARS CoV and anti-SARS-CoV-2 S-RBD IgG **(D):** Heat map from highest (top) to lowest (bottom) levels of anti-S-RBD Ig isotypes X marks missing samples. **(E):** Age negatively correlated with SARS-CoV-2 S-RBD IgG both at 1- and 6- months post vaccine; r=-0.413, p=0.029 (Group 1 only), r=-0.401, p=0.028 (all subjects) respectively; Pearson correlation. **(F)** Subjects in Groups 1 and 2 did not statistically differ in age p=0.44; Mann- Whitney test.

At baseline, during active wildfire exposure, elevated IL-13 expression in CD56^bright^ NK cells showed a positive correlation with total B cell counts, not only acutely, but also persisting at 1- and 6-months post- vaccination in Group 2 subjects (**Figure 3A**). Higher AQI levels on the day of vaccination of Group 1 subjects positively correlated with total IgG, 1-month post vaccine (**Figure 3B**). These data raise the possibility that pollutant exposure in association with elevated IL-13 expression in CD56^bright^ NK cells may drive non-specific immunoglobulin production and can have a lasting impact on non-specific B cell activation. In contrast, IL-13 expression in CD56^bright^ NK cells at the time of vaccination inversely correlated with S-RBD-specific IgG, 1-month post vaccine in the same Group 1 subjects, suggesting that excessive IL-13 production is associated with impaired quality of the antibody response to vaccination (**Figure 3C**). Further, Group 1 (vaccinated during wildfires) exhibited increased total B cells and significantly reduced S-RBD-specific IgG levels 6 months post-vaccination than Group 2 (vaccinated outside of wildfire periods) (**Figure 3E-F**). The 2-week average of AQI around the time of vaccination was negatively associated with S-RBD-specific IgG levels at 6 months in both groups (**Figure 3G**). These results indicate that in association with proinflammatory NK cell activation, wildfire smoke may impair adaptive immunity to vaccines.

**Figure 3.**
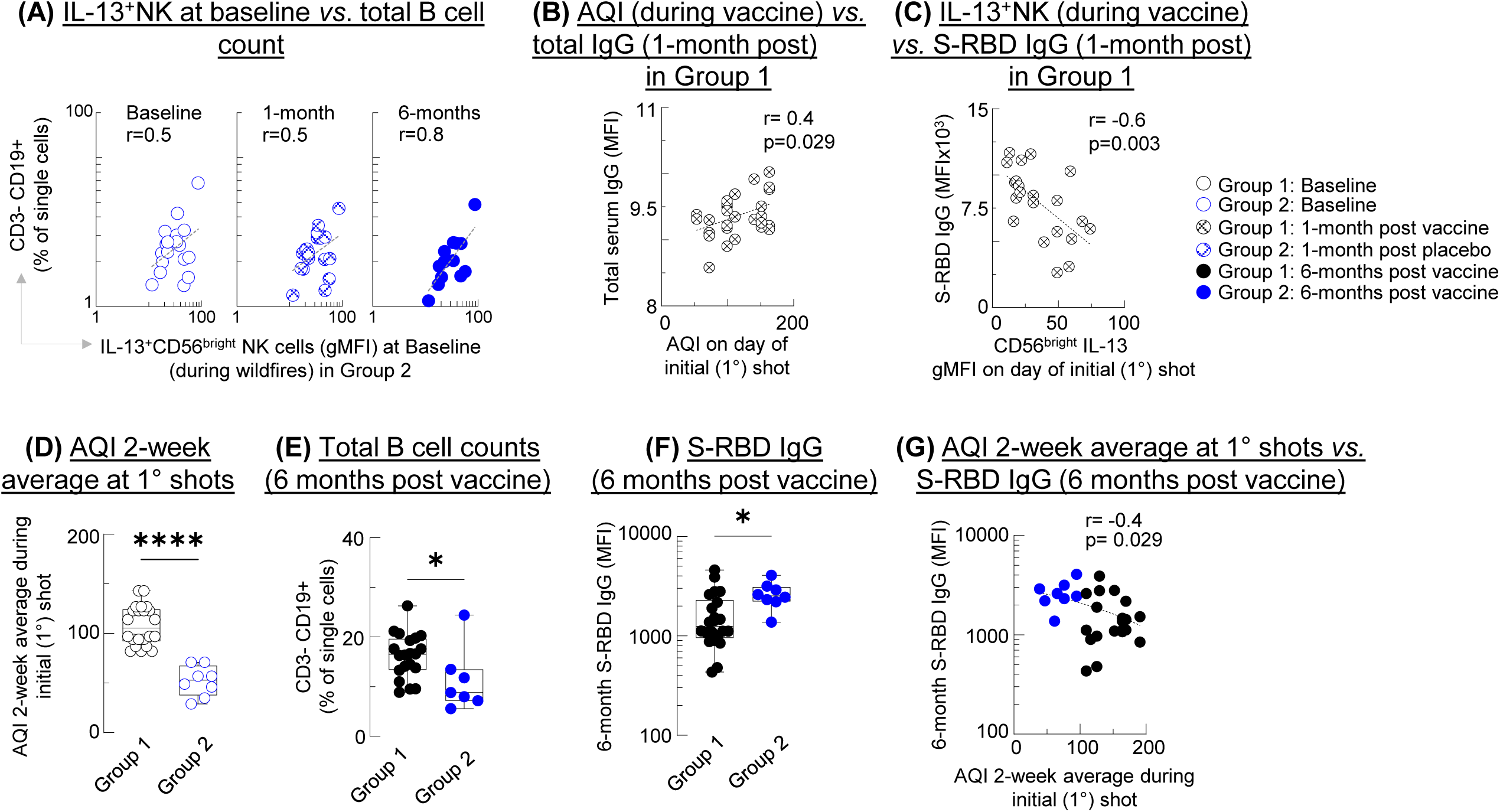
Wildfire smoke exposure and circulating NK cell expression of IL-13 correlated with impaired vaccine immunity. **(A):** CD56bright IL-13 at baseline (during wildfires) positively correlated with total B cell counts (% of singles) acutely, 1 month later and 6-months post vaccination. **(B):** AQI on day of initial shots positively correlated with total IgG in Group 1, 1-month post vaccination. **(C):** IL-13+CD56brightNK cells negatively correlated with S-RBD IgG in Group, 1-month post vaccination (Pearson correlation). **(D):** AQI 2-week average (1 week before and 1 week after) the day of initial vaccine shots. **(E-F):** Compared to Group 2 (vaccinated outside of wildfire period, blue dots), Group 1 (black dots) had and increased total (CD19+) B cells **(E)** and reduced S-RBD IgG **(F)** 6-months later. **(G)** AQI 2-week average during vaccination negatively correlated with S-RBD IgG 6 moths later. **(D):** Box and whiskers with interquartile ranges; **(D-F):** *p<0.05; ****p<0001; Mann-Whitney test (Group 1 vs. Group 2).

**Table 1.** Baseline demographics. Consented subjects were healthy, non-smoking, Sacramento Resident participants in the BNT162b2 COVID-19 vaccine trial at UC Davis Health.

|  | <u>**Group 1**</u><br>Vaccinated during<br>wildfires<br>**(N=28)** | <u>**Group 2**</u><br>Vaccinated outside of wildfire<br>periods<br>**(N=24)** |
| --- | --- | --- |
| **Age** (y), mean ± SD | 47.8 ± 14.4 | 45.7 ± 12.2 |
| **Sex:** male, n (%) | 15 (53.6%) | 16 (66.7%) |
| **BMI** (kg/m<sup>2</sup>), mean ± SD | 29.0 ± 5.8 | 30.8 ± 6.2 |
| **Ethnicity/Race** |  |  |
| Asian | 8 | 4 |
| American Indian, Alaska Native | 0 | 1 |
| Black, African American | 3 | 4 |
| Hispanic, LatinX | 3 | 3 |
| Native Hawaiian, Pacific Islander | 1 | 0 |
| White | 8 | 9 |
| Multiracial | 5 | 3 |

The real-world nature of our study while highly relevant also introduced limitations including the relatively small number of participants and the reliance on AQI as a proxy for smoke exposure. We found that age (not sex, BMI or race/ethnicity) negatively affected vaccine immunity but since the study was conducted in healthy individuals, other potentially important modifiers such as underlying health conditions were not investigated. While a few recent studies raised the general importance of air pollution in vaccine immunity especially against SARS-CoV-2^4, 5^, we uniquely conducted this work on the first round of vaccination against COVID-19 concurrent with wildfire smoke exposure. We suggest a previously unrecognized pathway involving wildfire-smoke induced CD56^bright^NK-cell IL-13, non-specific B activation and impaired vaccine immunity. These novel findings warrant future mechanistic and clinical studies and could inform strategies during vaccination campaigns especially for populations exposed to high levels of air pollution.

## Data Availability

All data produced in the present study are available upon reasonable request to the authors

## Acknowledgments

The authors would like to acknowledge the 52 study participants who donated blood samples and their time. We are also grateful for all the research coordinators who provided excellent help during the conduct of this study. This work was supported by T32 HL007013 (for GKS), the Chester Robbins Pulmonary Research Endowment and TRDRP 556126/2018A (both for AH)

## Abbreviations

AQI: Air Quality Index
BMI: Body Mass Index
FACS: Fluorescence-Activated Cell Sorting
IgG: Immunoglobulin G subclass
IL-13: Interleukin-13
NK: Natural Killer
O3: Ozone
PM2.5: Particulate Matter <2.5 µm
S-RBD: SARS-CoV-2 Spike Receptor Binding Domain

## ONLINE REPOSITORY

### Online Repository Methods

#### Study participants and samples

All participants provided informed consent as well as medical background information at the time of enrollment. Only healthy non-smoking individuals, or individuals with diseases that have resolved in the past, were recruited into this study. This study was approved by the UC Davis IRB.

52 healthy subjects from the Sacramento region (26-83 years old) who were recruited in the original Pfizer BioNTech BNT162b2 vaccine trial at UC Davis Medical Center in August 2020 consented to enroll into our study. Because of the real-world nature of our study (Figure 1A), we were originally blinded to the subjects’ group assignments. Subjects were administered either 30µg of the BNT162b2 vaccine or saline solution *via* two doses of intramuscular injection, approximately 21 days apart^4^. Once FDA approved the BNT162b2 vaccine (December 11, 2020), the group identities of the 240 Pfizer vaccine trial volunteers were revealed. Our study included 28 vaccinated (Group 1, black symbols throughout) and 24 placebo-injected (Group 2, blue symbols) individuals. The latter group received their vaccine several months later, outside of wildfire season, in March-April 2021. Thus, all 6-months samples were collected from subjects who were vaccinated with the Pfizer mRNA BNT162b2 COVID- 19 vaccine. The tick marks on the x axis in Figure 1A denote weeks.

We collected blood samples (20mL) by venipuncture in August 2020 (at *baseline*); in October 2020, (*1 month after vaccination/Group 1, or placebo/Group 2*);and then April-October 2021, (*6 months after vaccination*).

In Group 1 there were 28 samples at baseline and 1 month after vaccination and 18 samples at 6 months. In Group 2 there were 24 samples at baseline and 1 month and 8 samples at 6 months. Attrition was due to study drop-outs, COVID-19 infections (determined by PCR positivity, SARS-CoV-2 nucleocapsid presence or 6-months S-RBD IgG levels exceeding 1-month levels. In addition, to be able to compare 6-month Group 1 immunoglobulin levels (all collected outside of wildfire periods) with Group 2 levels, we had to remove 4 Group 2 samples collected during the Caldor fire under very high AQI (Figure 1B). That way the only difference was that Group 1 received the vaccine during wildfires while Group 2 received it outside of wildfires. Both groups were studied 6 months later outside of wildfires.

#### Air Quality Index (AQI) data collection and calculation

Figure 1A was prepared using Daily Air quality index (AQI) between August 1, 2020, and October 31, 2021, in Sacramento, CA, shown for fine particulate matter (PM2.5, dark red) and O3 (orange). The data points came from the United States Environmental Protection Agency (U.S. EPA) Air Data – Tile Plot ([Air Data - Tile Plot | US EPA](https://www.epa.gov/outdoor-air-quality-data/air-data-tile-plot)) by selecting: 1. Pollutant (PM2.5 and pzone); 2. Year (2020 and 2021); 3. Geographic Area (California, Sacramento County) and 4. Monitor Site (060670010). The colors for the six AQI categories are shown by the y axis and the EPA definitions are listed on the right. The higher the AQI value, the greater the level of air pollution. AQI values are calculated using NowCast for each pollutant originally measured in parts per million (ppm) for ozone and in µg/m^3^ for PM2.5.

The combined (PM2.5 and O3) individual AQI data used for statistical analysis were collected from the AirNow Interactive Map (epa.gov) by selecting for the specific date, then ozone and PM2.5 monitors, and selecting the Downtown Sacramento – T Street Site (Site ID: 060670010, Agency: California Air Resources Board). AirNow is a partnership of the U.S. Environmental Protection Agency, National Oceanic and Atmospheric Administration (NOAA), National Park Service, NASA, Centers for Disease Control, and tribal, state, and local air quality agencies.

In **Figure E1** we show that the daily AQI measurements (on four randomly selected dates) were comparable across 4 monitoring stations in Sacramento County suggesting that the study subjects were similarly exposed by wildfire smoke. Figure 1B shows the individual AQI values on the days blood samples were drawn from Group 1 and Group 2 at baseline; 1 month after vaccination (Group 1) or placebo injection (Group 2); and 6 months after vaccination in both groups. The bracket indicates the Group 2 6-month collections that occurred during the Caldor Fire. These samples were included into the correlation assessing acute wildfire smoke effects with NK cell IL-13 expression on the day of blood draw (shown in Figure 1D-E. However, to be able to compare 6-month immunoglobulin levels between Group 1 (which were taken outside of wildfire period) with Group 2, the samples obtained during the Caldor fire had to be removed from assessment.

#### Peripheral blood mononuclear cell isolation

20 mL of peripheral blood was collected by venipuncture. Blood was layered over Ficoll-Paque PLUS(Amersham Biosciences, Piscataway, NJ) and centrifuged at 20°C for 25 minutes, brake and acceleration speed set to 0. Cells were harvested from the interphase layer and washed twice with a solution of PBS and 2% FBS. Cells were counted via the Countess® Automated Cell Counter (Thermo Fisher Scientific, Waltham, MA) then apportioned for downstream flow cytometry experiments.

#### Flow cytometry

Fluorescent-conjugated monoclonal antibodies were purchased from Biolegend (San Diego, CA), BD Biosciences (San Jose, CA), or eBioscience (San Diego, CA). Single-cell suspensions of peripheral blood mononuclear cells were plated in 96-well round bottom plates. Cells were stained with surface markers in FACS buffer (PBS + 2% FBS) for 30 minutes. Cells were washed in FACS buffer and then fixed and permeabilized by eBioscience™ Foxp3/ Transcription Factor Staining Buffer Kit according to the manufacturer’s instructions. Cells were then stained for intracellular markers in FACS buffer for 30 minutes at room temperature, and analyzed by resuspending in FACS buffer and running them on LSR II Flow Cytometer (BD Biosciences, San Jose, CA). Data was generated using FACSDiva software and analyzed on FlowJo V10.8.1 (Tree Star, OR). Cells were stained with the following antibodies purchased from BD Biosciences: anti-CD56-Alexa Fluor 488 (clone B159) and anti-CD3-PCP-Cy5.5 (clone SP34-2), and the following antibodies purchased from Biolegend: anti-CD4-APC (clone A161A1), anti-CD8-APC-Cy7 (clone RPA-T8), anti-IL-13-PE (clone JES10-5A2) (Biolegend), anti-CD19-BV650 (clone HIB19), and anti-CD20-BV785 (clone 2H7).

Peripheral blood NK cells gated using standard strategy were stratified into CD56^bright^ and CD56^dim^ populations and IL-13 expression was assessed in both. To control for non-specific fluorescence, PE- GRA (gMFI) gated on B cells (normally no- or very low IL-13 expressors) was subtracted from every corresponding sample of every subject.

#### Quantifying antibodies against SARS-CoV-2

A multiplex test was developed at the UC Davis Medical Center, Sacramento, CA that measured antibodies against spike (S) and Nucleocapsid (N) proteins of SARS-CoV-2, as well as antibodies against SARS-CoV, MERS-CoV and common human coronavirus strains (229E, NL63, OC43, HKU1). Anti-N antibodies differentiated subjects who received the BNT162b2 vaccine from those who got naturally infected with SARS-CoV-2^16^.

#### Data Analysis

Statistical analysis was performed using Prism v9 software (GraphPad Inc., La Jolla, CA). Wilcoxon matched-pairs signed rank test was used to compare the effects of the vaccine and the wildfire. A p- value less than 0.05 was considered significant.

### Online Repository Figure Legends

**Figure E1: Measurements of daily AQI were comparable on four randomly selected dates across 4 monitoring stations (represented by the 4 different bars) in Sacramento County.**

**Figure E2: Anti-MERS CoV S-RBD, nucleocapsid (N) or total IgG levels were not induced by BNT162b2 COVID-19 vaccination (A):** Immunoglobulin levels were measured by LUMINEX before, and 1 and 6-months post vaccination for MERS CoV S-RBD IgG, SARS-CoV-2 nucleocapsid (N) and total IgG **(B):** Antibody levels against 4 common human coronaviruses were not affected.

**Figure E3: Sex, race, ethnicity or body mass index did not affect BNT162b2 vaccine immunity. (A):** There was no difference in S-RBD IgG levels between sexes. **(B):** Race/ethnicity did not affect S-RBD IgG levels; one-way ANOVA. **(C):** Body Mass Index (BMI) did not affect S-RBD IgG levels.

---

**Figure E1.**
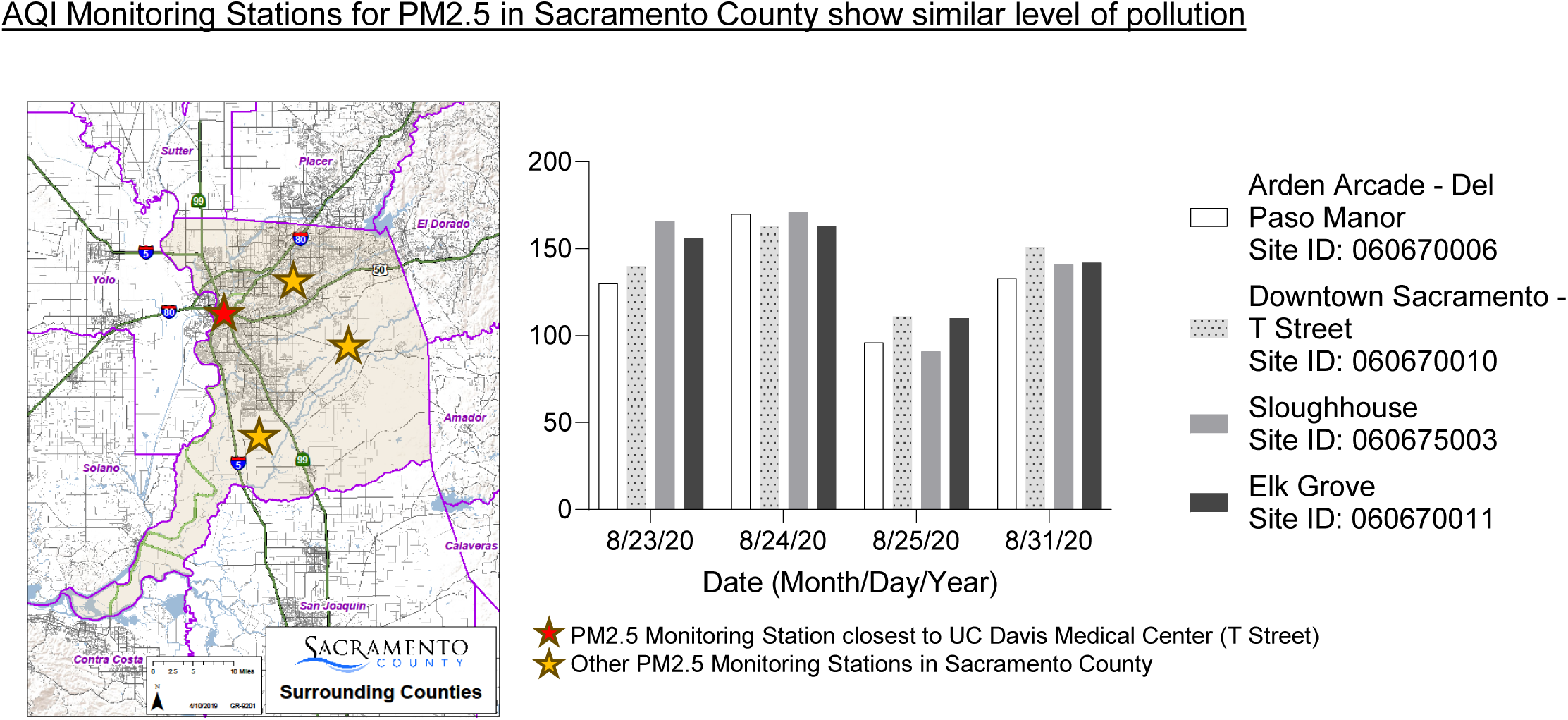
The daily AQI measurements were compared on four randomly selected dates across 4 monitoring stations (represented by the 4 different bars), in Sacramento County.

**Figure E2.**
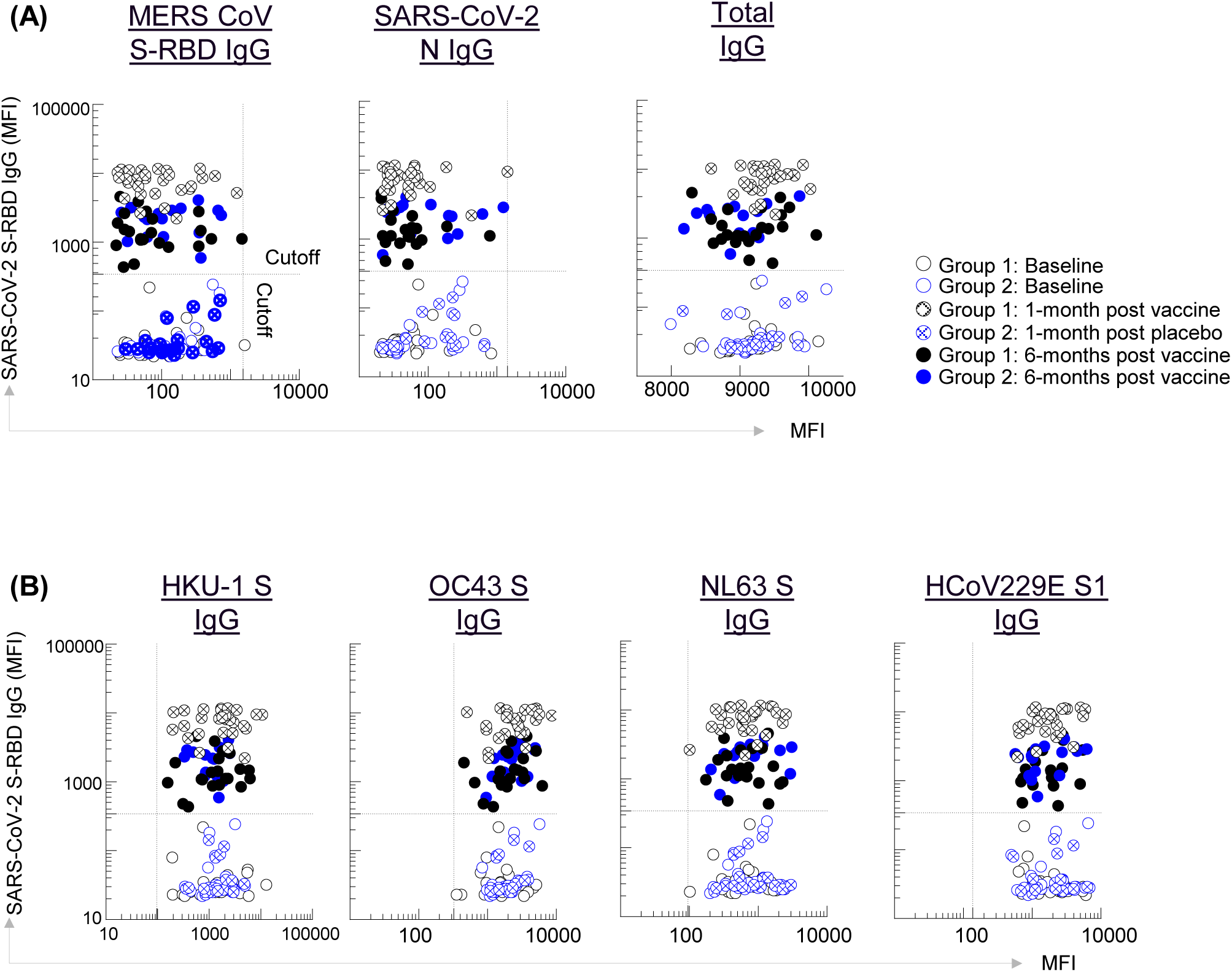
Anti-MERS CoV S-RBD, nucleocapsid (N) or total IgG levels were not induced by BNT162b2 COVID-19 vaccination. **(A):** Immunoglobulin levels were measured by LUMINEX before and 1 and 6-months post vaccination for MERS CoV S-RBD IgG, SARS-CoV-2 nucleocapsid (N) and total IgG **(B):** Antibody levels against 4 common human coronaviruses were not affected.

**Figure E3.**
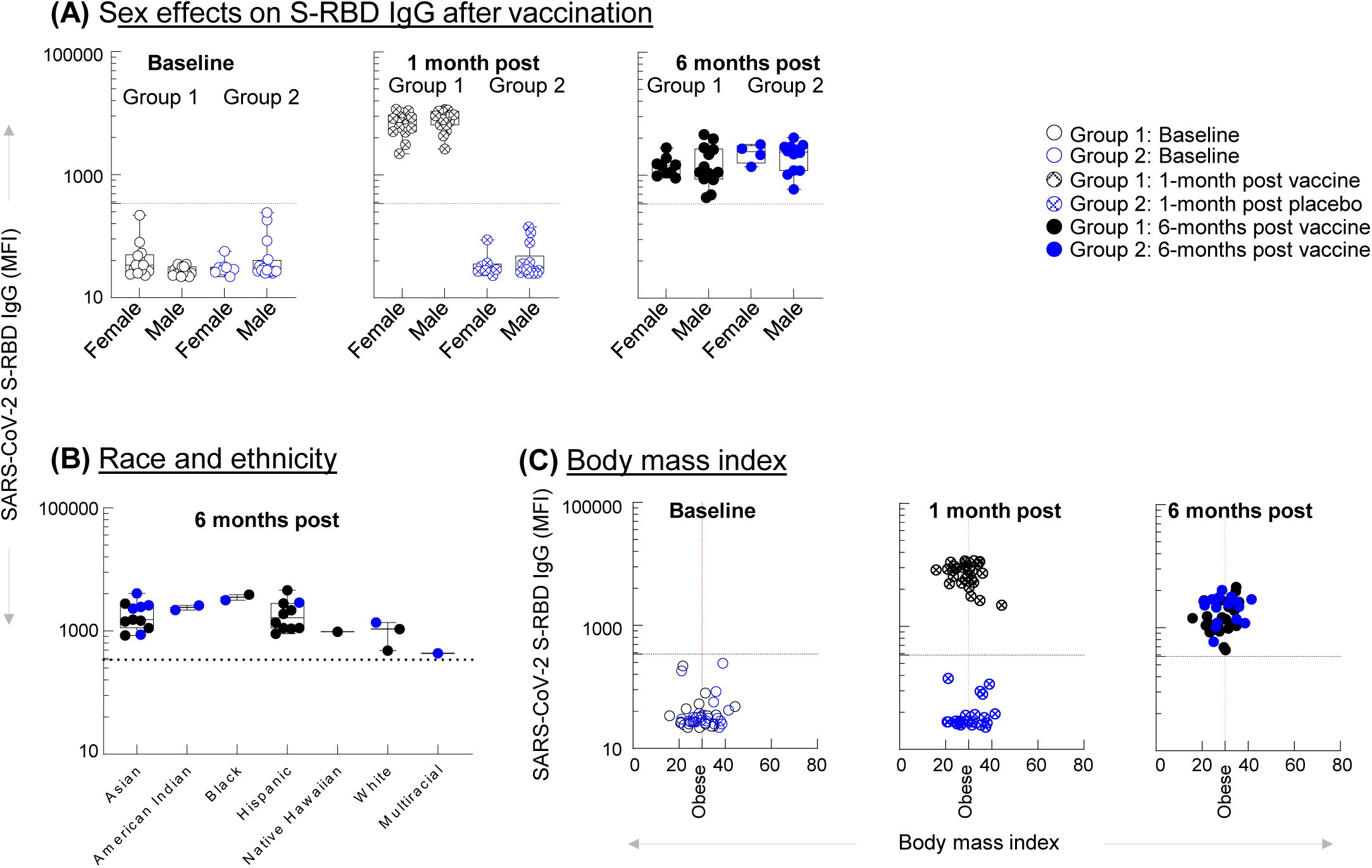
Determinants of BNT162b2 vaccine immunity. **(A)**: There was no difference in S-RBD IgG levels between sexes. **(B):** Race/ethnicity did not affect S-RBD IgG levels; one-way ANOVA. **(C):** Body Mass Index (BMI) did not affect S-RBD IgG levels.

## References

1. Bowman WS, Schmidt RJ, Sanghar GK, Thompson Iii GR, Ji H, Zeki AA, et al. “Air That Once Was Breath” Part 2: Wildfire Smoke and Airway Disease - “Climate Change, Allergy and Immunology” Special IAAI Article Collection: Collegium Internationale Allergologicum Update 2023. Int Arch Allergy Immunol 2024; 185:617–30.

2. Weheba A, Vertigan A, Abdelsayad A, Tarlo SM. Respiratory Diseases Associated With Wildfire Exposure in Outdoor Workers. J Allergy Clin Immunol Pract 2024; 12:1989–96.

3. Hertelendy AJ, Howard C, Sorensen C, Ranse J, Eboreime E, Henderson S, et al. Seasons of smoke and fire: preparing health systems for improved performance before, during, and after wildfires. Lancet Planet Health 2024; 8:e588–e602.

4. Kogevinas M, Karachaliou M, Espinosa A, Aguilar R, Castano-Vinyals G, Garcia-Aymerich J, et al. Long-Term Exposure to Air Pollution and COVID-19 Vaccine Antibody Response in a General Population Cohort (COVICAT Study, Catalonia). Environ Health Perspect 2023; 131:47001.

5. Kogevinas M, Castano-Vinyals G, Karachaliou M, Espinosa A, de Cid R, Garcia-Aymerich J, et al. Ambient Air Pollution in Relation to SARS-CoV-2 Infection, Antibody Response, and COVID- 19 Disease: A Cohort Study in Catalonia, Spain (COVICAT Study). Environ Health Perspect 2021; 129:;.

6. Ural BB, Caron DP, Dogra P, Wells SB, Szabo PA, Granot T, et al. Inhaled particulate accumulation with age impairs immune function and architecture in human lung lymph nodes. Nat Med 2022; 28:2622–32.

7. Bjorkstrom NK, Strunz B, Ljunggren HG. Natural killer cells in antiviral immunity. Nat Rev Immunol 2022; 22:112–23.

8. Karimi K, Forsythe P. Natural killer cells in asthma. Front Immunol 2013; 4:159.

9. Michel JJ, Griffin P, Vallejo AN. Functionally Diverse NK-Like T Cells Are Effectors and Predictors of Successful Aging. Front Immunol 2016; 7:530.

10. Ge MQ, Kokalari B, Flayer CH, Killingbeck SS, Redai IG, MacFarlane AWt, et al. Cutting Edge: Role of NK Cells and Surfactant Protein D in Dendritic Cell Lymph Node Homing: Effects of Ozone Exposure. J Immunol 2016; 196:553–7.

11. Graydon EK, Conner TL, Dunham K, Olsen C, Goguet E, Coggins SA, et al. Natural killer cells and BNT162b2 mRNA vaccine reactogenicity and durability. Front Immunol 2023; 14:1225025.

12. Rydyznski CE, Waggoner SN. Boosting vaccine efficacy the natural (killer) way. Trends Immunol 2015; 36:536–46.

13. Polack FP, Thomas SJ, Kitchin N, Absalon J, Gurtman A, Lockhart S, et al. Safety and Efficacy of the BNT162b2 mRNA Covid-19 Vaccine. N Engl J Med 2020; 383:2603–15.

14. Qian Q, Chowdhury BP, Sun Z, Lenberg J, Alam R, Vivier E, et al. Maternal diesel particle exposure promotes offspring asthma through NK cell-derived granzyme B. J Clin Invest 2020; 130:4133–51.

15. Aguilera J, Kaushik A, Cauwenberghs N, Heider A, Ogulur I, Yazici D, et al. Granzymes, IL-16, and poly(ADP-ribose) polymerase 1 increase during wildfire smoke exposure. J Allergy Clin Immunol Glob 2023; 2.

16. Ravindran R, Kang H, McReynolds C, Sanghar GK, Chang WLW, Ramasamy S, et al. Dynamics of temporal immune responses in nonhuman primates and humans immunized with COVID-19 vaccines. PLoS One 2023; 18:e0287377.

